# Shared Decision-Making in Renal Replacement Therapy Selection: Patient Perceptions, Preferences, and Influencing Factors in a Nationwide Cross-Sectional Study in Japan

**DOI:** 10.1101/2025.01.11.25320378

**Authors:** Yugo Shibagaki, Tadashi Sofue, Hiroo Kawarazaki, Tatsunori Toida, Tomo Suzuki, Hiroki Nishiwaki, Kenichiro Asano, Hiroyuki Terawaki, Takafumi Ito, Hideaki Oka, Kei Nagai, Minoru Murakami, Kojiro Nagai, Daisuke Komukai, Takayuki Adachi, Satoshi Furukata, Takaaki Tsutsui, Kiichiro Fujisaki, Seita Sugitani, Hideaki Shimizu, Tomoya Nishino, Hiroaki Asada, Hideki Shimizu, Tatsuo Tsukamoto, Izaya Nakaya, Yosuke Yamada, Ryohei Inanaga, Shohei Yamada, Shohei Nakanishi, Atsuhiro Maeda, Mari Yamamoto, Shuma Hirashio, Takeshi Okamoto, Takayuki Nakamura, Ken-ichi Miyoshi, Hiroshi Kado, Susumu Toda, Shigeru Shibata, Keiko Nishi, Makoto Yamamoto, Tsukasa Naganuma, Ryo Zamami, Masahide Furusho, Hitoshi Miyasato, Yukihiro Tamura, Yoshihiko Raita, Chisato Fukuhara, Keita Uehara, Kosuke Inoue, Yasuhiro Taki, Nobuyuki Nakano, Noriaki Kurita, the PREPARES Study Group

## Abstract

**Background:** Shared decision-making (SDM) is a key process in selecting renal replacement therapy (RRT). This study analyzed SDM perceptions, preferences, and patient- and facility-level factors influencing SDM among Japanese patients with chronic kidney disease (CKD) who selected RRT.

**Methods:** We analyzed 475 adult patients with CKD from 49 medical facilities. SDM awareness and recognition, preferences for SDM timing and frequency, discussion content, and desired professional involvement were assessed. Patient- and facility-level factors associated with SDM perceptions were evaluated using Poisson regression with cluster-robust variance estimation.

**Results:** The mean age of participants was 67.4 years. Overall, 71%, 24.4%, and 4.4% chose hemodialysis, peritoneal dialysis, and kidney transplantation, respectively. While 81.2% perceived that SDM was performed during RRT selection, only 4.7% were explicitly aware of the concept. Patients prioritized discussions about effects on daily life, financial burden, and family-related concerns. Most preferred SDM initiation when RRT was imminent, conducted over multiple sessions. Many patients valued the involvement of medical social workers and their usual non-nephrologist physicians in addition to nephrologists. Multiple outpatient visits for RRT selection, involving nurse participation and extended consultation times, were significantly associated with SDM perceptions (prevalence ratio: 1.59, 95% confidence interval: 1.05–2.42).

**Conclusions:** Many Japanese patients with CKD perceived SDM during RRT selection; however, they had limited awareness of the concept. The findings underscore the importance of establishing a system that facilitates repeated SDM discussions at critical moments for patients. These discussions should emphasize the impact of RRT on patients’ lives and involve a multidisciplinary team.

## Introduction

The decision-making process for patients with chronic kidney disease (CKD) to select a renal replacement therapy (RRT) modality, such as hemodialysis, peritoneal dialysis, or renal transplantation, is a critical issue. A transition from informed choice to shared decision-making (SDM) has been recommended, as each RRT option is considered a reasonable alternative depending on the patient’s circumstances.^1^ SDM is a collaborative approach in which physicians and patients work together to choose an RRT modality, with the patient contributing expertise on their individual circumstances, values, and goals, and the physician providing expertise on the disease and the risks and benefits of the available treatments.^2^ SDM can reduce regret associated with RRT selection^3^ and may improve treatment adherence,^4^ reduce emergency hospitalizations, and enhance overall prognosis.^5,6^ However, the optimal timing for initiating SDM, content of discussions, appropriate participants, and factors influencing RRT selection via SDM remain poorly understood.

First, patients with CKD and their physicians face significant challenges in deciding the timing and scope for discussing the choice of RRT.^2^ When kidney failure is not imminent, patients may perceive RRT as a distant concern.^2^ Second, patients often prioritize the impact of RRT on daily life—such as social participation and employment—over life-sustaining benefits and risks associated with the therapy.^7^ Third, decision-making participants may extend beyond the patients themselves and their nephrologists. While patient autonomy is emphasized in the United States and Europe, with family involvement viewed as either beneficial or detrimental to RRT decision-making,^3,8^ in Asia, including Japan, family involvement is often considered indispensable.^4,9^ Additionally, some patients seek advice from non-nephrologist healthcare providers with whom they have established long-term relationships.^2^ Fourth, while most discussions on promoting SDM focus on patients and healthcare providers, there is growing recognition of the need to address system-level factors, such as the involvement of multidisciplinary healthcare teams and the medical reimbursement system.^8^ For instance, the Japanese government reimburses outpatient visits for RRT selection if certain facility requirements are met; however, we do not know whether this policy facilitates SDM implementation.

Clarifying these aspects is essential for advancing SDM implementation. Therefore, we conducted a nationwide cross-sectional study involving Japanese adults with CKD who had chosen an RRT modality but had not yet initiated treatment.

## Materials and Methods

### Design, setting, and participants

This multicenter cross-sectional study was conducted between October 2022 and September 2024 at 49 facilities across Japan providing outpatient care for patients with CKD, including dialysis initiation and, at some sites, renal transplantation (Supplementary Figure S1). The study sites included affiliated facilities where alumni of the primary investigator’s department practice, nearby facilities in the Kanto, Shikoku, and Kyushu regions where co-investigators work, and other facilities with participating nephrologists connected to the research team. These facilities were invited to participate based on personal networks within the nephrology community. The eligibility criteria were: (1) adult patients with stage 5 CKD, (2) those who had selected one of the modalities of RRT, and (3) those who had not yet initiated that RRT. Exclusion criteria were (1) patients in whom RRT was emergently initiated before the choice of RRT was made, (2) patients who were too impaired in physical or cognitive function to complete the questionnaire, or (3) patients who chose conservative kidney management. Patients were rewarded with a 500-yen gift card for completing the questionnaire. The study was approved by the Fukushima Medical University Certified Review Board, Fukushima Medical University (No. ippan-2022).

### The questionnaire regarding the choice of RRT and SDM

The self-administered questionnaire, detailed in Supplementary Items S2 to S4, was developed by the study authors. This questionnaire included items on participant demographics (e.g., education, income, employment status), chosen RRT modality, medical conditions discussed for selection of and preparation for RRT, foundational knowledge on SDM, preferred healthcare provider involvement in SDM, desired information for SDM, and the timing and frequency of discussion using SDM. Both paper and digital versions of the questionnaire were offered, with digital responses enabled via a QR code link on the paper form. Participants were assured that their responses would remain confidential and would not be reviewed by their attending physicians. This policy was upheld through oversight by principal investigators at each site. Responses submitted via paper were directed to a centralized analysis facility for processing and data integration.

### Collecting clinical and facility data

Facility characteristics and clinical data were collected from treating physicians. Patient data included age, sex, primary cause of end-stage kidney disease (ESKD), frailty (as assessed by the Japanese version of the Clinical Frailty Scale, version 2.0^10,11^), and the number of outpatient visits for RRT selection. Outpatient visits for RRT selection required the involvement of nursing staff and a minimum consultation time of 30 minutes. However, these visits did not need to be separate from regular nephrology follow-up appointments. Additionally, the facility providing these services was not required to meet specific criteria to claim teaching and management fees for RRT under Japan’s unique reimbursement system, which allows up to two claims per patient. For example, the facility did not need to have a prior history of claiming reimbursement for peritoneal dialysis teaching and management.

The facility characteristics included the total number of beds, the typical annual volume of RRT initiations, the actual volume of RRT initiations in the most recent quarter, the availability of specialized outpatient visits for RRT selection, the frequency of multi-professional healthcare provider involvement in RRT selection discussions, and the types of physicians participating in this process.

### Statistical analysis

All statistical analyses were performed using Stata/SE version 18. For summarizing patient characteristics, facility characteristics, and responses regarding SDM, continuous variables were described as means and standard deviations, and categorical variables were described as frequencies and percentages.

Sankey diagrams were created to visually represent the distribution of the combined frequency of SDM initiation timing and implementation frequency before RRT selection, as well as the combined frequency of SDM implementation frequency after RRT selection but prior to initiation, and the willingness to engage in SDM following RRT initiation.^12^

The potential factors associated with strong agreement with having the choice of RRT through the SDM approach were studied via exploratory analysis. For this, we fit a Poisson regression with cluster-robust variance estimation with facilities as cluster units to estimate the prevalence ratio of the strong agreement.^13^ This rationale is based on the fact that, unlike odds ratios, the ratio of proportions of strong agreement can be estimated without overestimating the ratio when strong agreement is not rare. Candidate predictors were chosen based on our clinical expertise and forced into the model. For any predictor with missing values, multiple imputation with chained equations was performed assuming that the missing values were at random. Estimates from 10 imputed data were combined into a single estimate.

## Results

### Participant Characteristics

The patient (N = 475) characteristics are summarized in Table 1. The mean age was 67.4 years (SD, 13.1), 4–5 years younger than the average age of patients undergoing incident dialysis in Japan, based on data from the Japanese Society for Dialysis Therapy (JSDT) registry as of 2022.^14^ Overall, 65.3% of participants were male, and the leading causes of ESKD were diabetic kidney disease (38.1%), nephrosclerosis (21.1%), and chronic glomerulonephritis (19.8%), and sex distribution, and primary causes of ESKD were comparable with those of the general Japanese incident dialysis population.^14^ More than half of the participants had an education level of high school or below (62.1%) and an annual household income below 5 million yen (63.8%). Nearly 60% were retired or unemployed, while 38.4% were currently working. Based on the clinical frailty scale, approximately 20% of participants were classified as having frailty. Regarding RRT modality, 71.0%, 24.4%, and 4.4% of participants selected hemodialysis, peritoneal dialysis, and kidney transplantation, respectively, suggesting a higher rate of non-hemodialysis selection than the national average.^14^ Overall, 44.2% of patients had a single outpatient visit for RRT selection, while 30.3% attended two or more visits.

**Table 1.** Patient characteristics.

| **Patient characteristics, N = 475** |  |
| --- | --- |
| **Age, year** | 67.4 (13.1) |
| **Male sex, n (%)** | 310 (65.3) |
| **Cause of ESKD, n (%)** |  |
| Diabetic kidney disease | 181 (38.1) |
| Chronic glomerulonephritis | 94 (19.8) |
| Nephrosclerosis | 100 (21.1) |
| Polycystic kidney disease | 23 (4.8) |
| Others/Unknown | 77 (16.2) |
| **Education level, n (%)** |  |
| Junior high school graduate or less | 73 (15.7) |
| High school graduate | 216 (46.4) |
| University/Graduate school graduate | 81 (17.4) |
| Others | 96 (20.6) |
| *missing, n* | 9 |
| **Household income, n (%)** |  |
| <1,000,000 yen | 39 (8.8) |
| 1,000,000–<5,000,000 yen | 284 (63.8) |
| 5,000,000–<10,000,000 yen | 86 (19.3) |
| >10,000,000 yen | 36 (8.1) |
| *missing, n* | 30 |
| **Working, n (%)** |  |
| Yes | 181 (38.4) |
| On leave | 14 (3.0) |
| No | 277 (58.7) |
| *missing, n* | 3 |
| **Frailty, n (%)** |  |
| No frailty | 379 (79.8) |
| Very mild/Mild frailty | 76 (16.0) |
| Moderate/Severe frailty | 20 (4.2) |
| Very severe frailty/Terminal ill | (0) |
| **No. of outpatient visit for RRT selection\*, n (%)** |  |
| None | 121 (25.5) |
| Once | 210 (44.2) |
| Twice ore more | 144 (30.3) |
| **Engagement of primary nephrologist in RRT selection, n (%)** |  |
| Yes | 394 (86.2) |
| No | 63 (13.8) |
| *missing, n* | 18 |
| **Selected RRT, n (%)** |  |
| Hemodialysis | 337 (71.0) |
| Peritoneal dialysis | 116 (24.4) |
| Kidney transplantation | 21 (4.4) |
| Hemodialysis and peritoneal dialysis | 1 (0.2) |
| **Discussions with their physicians treating kidney disease\*\*, n (%)** |  |
| To do or not to pursue RRT | 368 (79.3) |
| Which type of RRT to choose | 373 (80.4) |
| To proceed with or not yet proceed with medical preparation for the selected RRT | 359 (77.4) |
| To start or not yet start the selected RRT in the near future | 347 (74.8) |
| *missing, n* | 11 |
| Continuous variables are summarized as the mean with the standard deviation in parentheses, whereas categorical variables are presented as the frequency with the percentage in parentheses. |  |
| *Outpatient visits for RRT selection (involving nursing staff collaboration and requiring a minimum of 30 min) were neither required to meet facility criteria for reimbursement claims nor mandated to be separate from regular nephrology outpatient visits. |  |
| **Multiple choice allowed. |  |
| RRT, renal replacement therapy; ESKD, end-stage kidney disease |  |

### Facility Characteristics

The participating facilities varied in bed capacity: 4.1% were clinics, and 8.2%, 18.4%, and 69.4% were low-, low-to-moderate, and moderate-to-high capacity hospitals (Table 2). Specialized outpatient visits for RRT selection were available at 65.3% of the facilities. Regarding multi-professional participation in explaining RRT options, two-thirds of facilities (67.4%) reported that multi-professional teams were not involved 70–80% of the time. For physician involvement, in 87.8% of the facilities, the patient’s usual physician was responsible for explaining RRT options most of the time (70–80% of the time).

**Table 2.** Facility characteristics.

| **Facility characteristics, N = 49** |  |
| --- | --- |
| **Facility type by bed capacity, n (%)** |  |
| Clinic (No. of beds <20) | 2 (4.1) |
| Low-capacity hospital (No. of beds 20–<200) | 4 (8.2) |
| Low-to-moderate capacity hospital (No. of beds 200–<400) | 9 (18.4) |
| Moderate-to-high capacity hospital (No. of beds ≥400) | 34 (69.4) |
| **Typical annual volume of RRT initiations, n (%)** |  |
| <30 cases | 15 (30.6) |
| 30–100 cases | 26 (53.1) |
| >100 cases | 8 (16.3) |
| **Actual volume of RRT initiations in the most recent quarter, cases\*** | 14.1 (9.3) |
| **Specialized outpatient visit for RRT selection, n (%)** |  |
| Absent | 17 (34.7) |
| Present | 32 (65.3) |
| **Multi-professional participation in explaining choice of RRT, n (%)** |  |
| Most of the time (70–80% of the time), yes | 9 (18.4) |
| Most of the time (70–80% of the time), no | 33 (67.4) |
| Vary depending on the case | 2 (4.1) |
| Outpatient service in choice of RRT provided by nurses only | 5 (10.2) |
| **Physicians explaining choice of RRT, n (%)** |  |
| Most of the time (70–80% of the time), the patient's usual nephrologist | 43 (87.8) |
| Most of the time (70–80% of the time), the physician in charge of specialized outpatient visits for RRT selection | 5 (10.2) |
| Vary depending on the case | 1 (2.0) |
| Continuous variables are summarized as the mean with the standard deviation in parentheses, whereas categorical variables are presented as the frequency with the percentage in parentheses. |  |

\*The median was 12, with a range of 1 to 46.
RRT, renal replacement therapy

### Patient response to items regarding SDM

#### Awareness of SDM and perception of SDM process for RRT selection

Familiarity with SDM varied considerably (Table 3). Only a small proportion (4.7%) reported being well aware of SDM, with an additional 18.4% somewhat aware of the approach. A majority (57.7%) were completely unaware of the approach. A substantial proportion indicated a positive perception regarding SDM process for RRT selection, with 39.1% strongly agreeing and 42.1% somewhat agreeing.

**Table 3.**
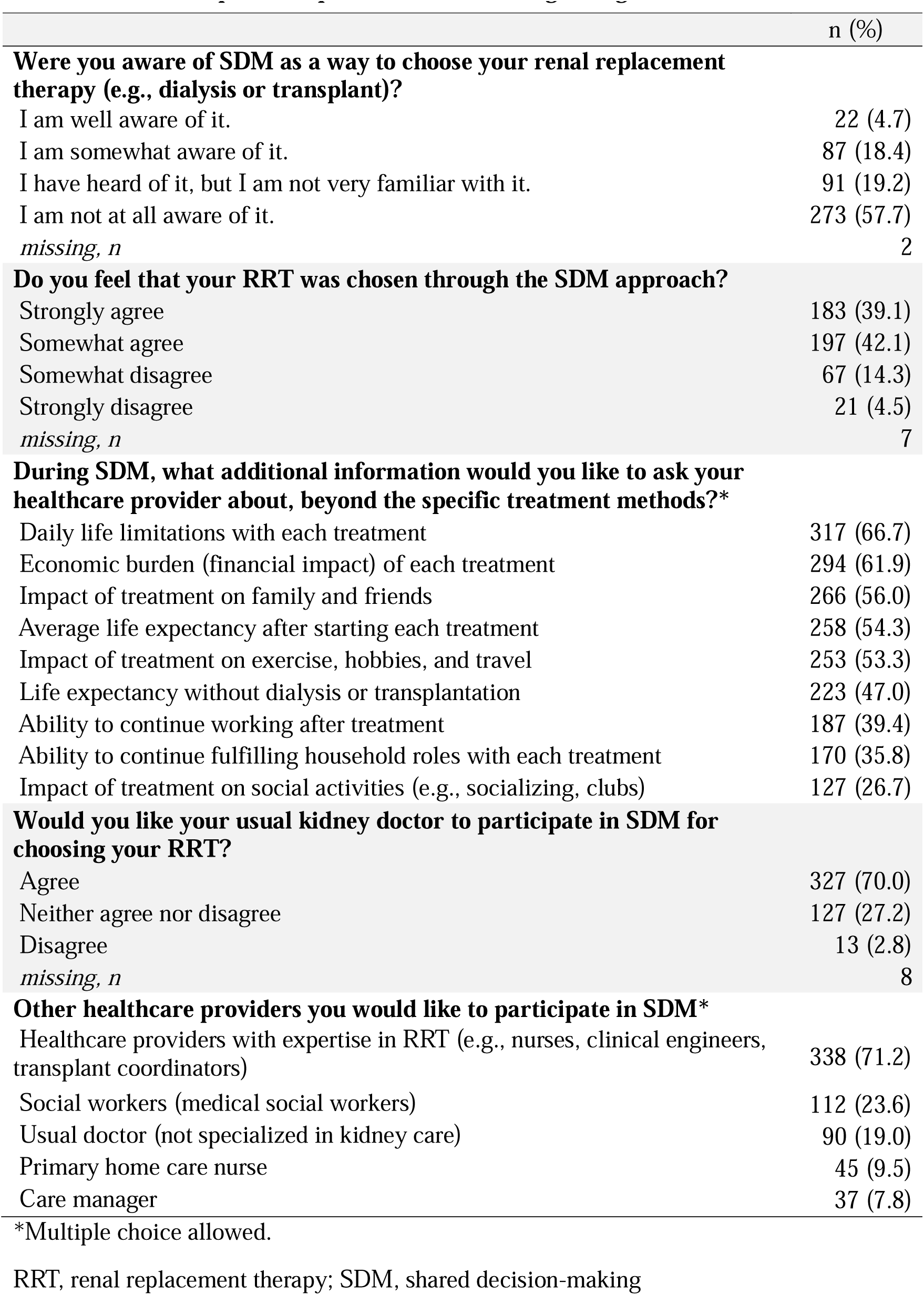
Patient response to questionnaire items regarding SDM.

|  | n (%) |
| --- | --- |
| **Were you aware of SDM as a way to choose your renal replacement therapy (e.g., dialysis or transplant)?** |  |
| I am well aware of it. | 22 (4.7) |
| I am somewhat aware of it. | 87 (18.4) |
| I have heard of it, but I am not very familiar with it. | 91 (19.2) |
| I am not at all aware of it. | 273 (57.7) |
| *missing, n* | 2 |
| **Do you feel that your RRT was chosen through the SDM approach?** |  |
| Strongly agree | 183 (39.1) |
| Somewhat agree | 197 (42.1) |
| Somewhat disagree | 67 (14.3) |
| Strongly disagree | 21 (4.5) |
| *missing, n* | 7 |
| **During SDM, what additional information would you like to ask your healthcare provider about, beyond the specific treatment methods?\*** |  |
| Daily life limitations with each treatment | 317 (66.7) |
| Economic burden (financial impact) of each treatment | 294 (61.9) |
| Impact of treatment on family and friends | 266 (56.0) |
| Average life expectancy after starting each treatment | 258 (54.3) |
| Impact of treatment on exercise, hobbies, and travel | 253 (53.3) |
| Life expectancy without dialysis or transplantation | 223 (47.0) |
| Ability to continue working after treatment | 187 (39.4) |
| Ability to continue fulfilling household roles with each treatment | 170 (35.8) |
| Impact of treatment on social activities (e.g., socializing, clubs) | 127 (26.7) |
| **Would you like your usual kidney doctor to participate in SDM for choosing your RRT?** |  |
| Agree | 327 (70.0) |
| Neither agree nor disagree | 127 (27.2) |
| Disagree | 13 (2.8) |
| *missing, n* | 8 |
| **Other healthcare providers you would like to participate in SDM\*** |  |
| Healthcare providers with expertise in RRT (e.g., nurses, clinical engineers, transplant coordinators) | 338 (71.2) |
| Social workers (medical social workers) | 112 (23.6) |
| Usual doctor (not specialized in kidney care) | 90 (19.0) |
| Primary home care nurse | 45 (9.5) |
| Care manager | 37 (7.8) |

\*Multiple choice allowed.
RRT, renal replacement therapy; SDM, shared decision-making

#### Additional Information Desired During SDM

Patients expressed interest in discussing a range of factors beyond specific treatment details. These most frequently included daily living restrictions (66.7%) and financial burdens associated with each treatment (61.9%), followed by information regarding the burden on family and friends (56%). In contrast, life expectancy after treatment initiation (54.3%) and life expectancy without dialysis or transplantation (47%) were less frequently cited.

#### Desire for the participation of physicians and other healthcare providers in SDM

Most (70%) patients wanted their treating physician to participate actively in the SDM. Patients also expressed a desire for various healthcare professionals knowledgeable about RRT, such as nurses, clinical engineers, and transplant coordinators (71.2%) to participate in SDM. The participation of their usual physicians outside of kidney care were also desired by 23.6% of respondents, along with medical social workers (19%), home care nurses (9.5%), and care managers (7.8%).

### Desired timing and frequency of SDM before RRT selection

The combination of timing and frequency shows that patients who desired earlier SDM initiation, such as 1–3 years before RRT initiation, opted for more frequent engagement before RRT selection, with many opting for “every visit” or “every few months” (Figure 1A). Conversely, patients who desired SDM within 6 months of RRT initiation chose less frequent engagement, such as “once” or “as needed.”

**Figure 1.**
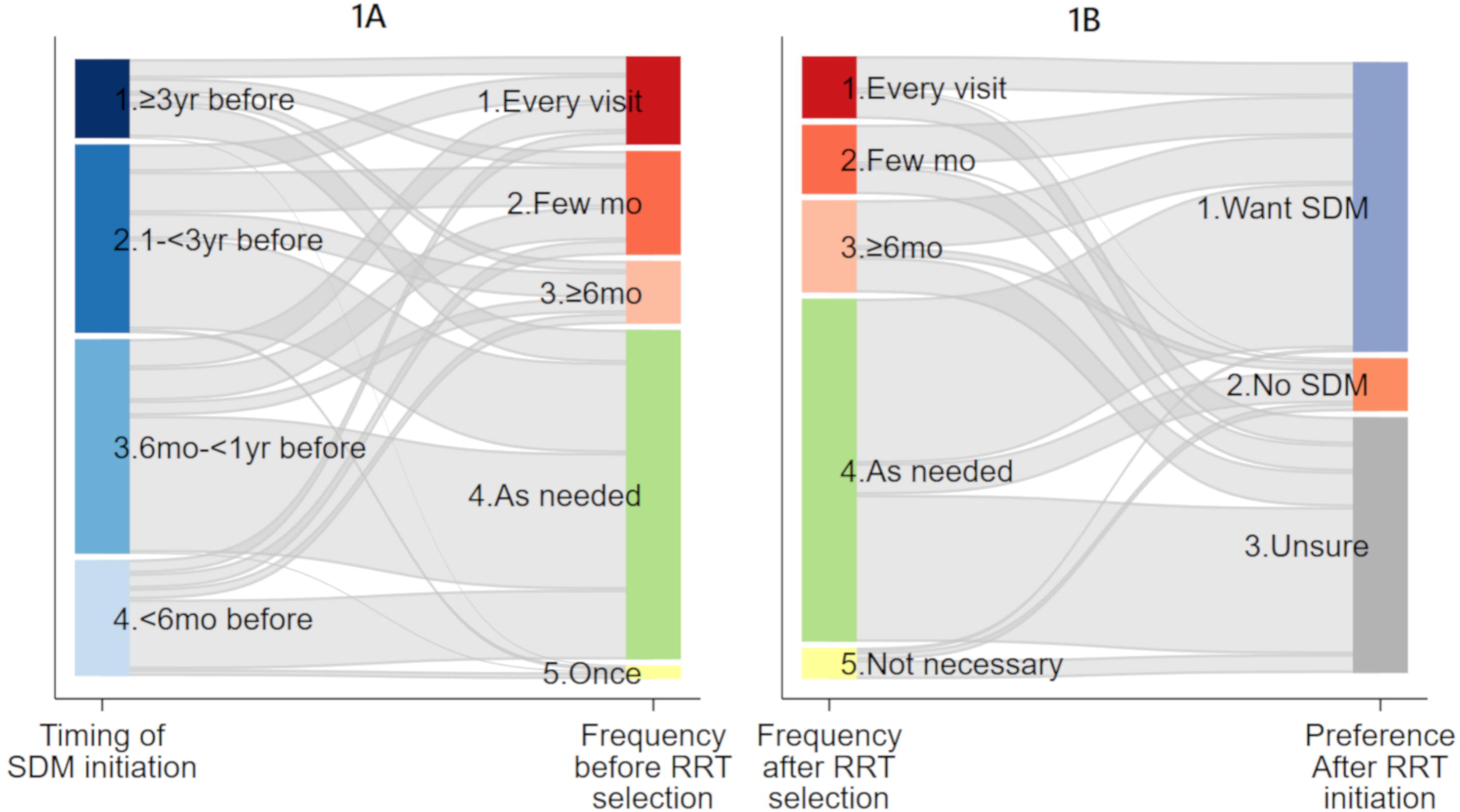
Sankey diagrams. 1A: A Sankey diagram illustrating the combination (flow) of the timing and frequency of SDM implementation before RRT selection (n = 438). The height of the individual boxes (nodes) on the vertical axis represents relative proportions, while the thickness of the links connecting the boxes for timing and frequency reflects the relative proportions of each combination. Deep blue indicates “More than 3 years before RRT”, medium blue indicates “1 to less than 3 years before RRT”, light blue indicates “6 months to less than 1 year before RRT”, and very light blue indicates” Less than 6 months before RRT”. Red indicates “Every visit,” orange indicates “Every few months,” light orange indicates “Every 6 months or more,” green indicates “As needed,” and light yellow indicates “Once.” For example, 3.2% of patients reported timing of “More than 3 years before” with a frequency of “Every visit.” In contrast, 1.1% reported timing of “6 months to less than 1 year before” with a frequency of “As needed,” and 22.8% reported timing of “Less than 6 months before” with a frequency of “Once.” 1B: A Sankey diagram illustrating the frequency of SDM implementation after RRT selection and preferences for SDM after RRT initiation (n = 440). The thickness of the links connecting the boxes for frequency of implementation and preferences after RRT initiation indicates the relative proportions of each combination. Red indicates “Every visit,” orange indicates “Every few months,” light orange indicates “Every 6 months or more,” green indicates “As needed,” and light yellow indicates “Not necessary.” Blue indicates “Want SDM,” orange indicates “No SDM,” and grey indicates “Unsure.”

### Desired frequency of SDM after RRT selection and preference for SDM after RRT initiation

Many patients who desired regular SDM after RRT selection (categorized as “Every” or “Few months”) expressed a continued desire for SDM (“Want SDM”) following RRT initiation (Figure 1B). Conversely, patients with less frequent (e.g., “as needed”) or no desire for SDM after RRT selection often reported “Unsure” or expressed no desire for further engagement in SDM (“No SDM”) after RRT initiation.

### Factors associated with a strong patient perception that RRT selection was via SDM

Several patient-level characteristics were associated with a strong perception that RRT selection was via SDM (Table 4). Compared with patients with an education level of junior high school or lower, those with a high school education (prevalence ratio [PR]: 0.61, 95% CI: 0.44–0.83), those with a university or graduate school education (PR: 0.64, 95% CI: 0.42–0.95) and those in the “other” education category (PR: 0.75, 95% CI: 0.57–0.996) were less likely to perceive RRT selection as involving SDM. Patients with an annual income of less than 1,000,000 yen were less likely to perceive RRT selection as involving SDM than those with an income of over 10,000,000 yen (PR: 0.53, 95% CI: 0.32 to 0.86). Patients who had two or more outpatient visits for RRT selection were more likely to report perceived SDM than those who had no visits (PR: 1.59, 95% CI: 1.05 to 2.42).

**Table 4.** Associations of perceived SDM with patients’ and facilities’ covariates† (n = 468)

|  | Adjusted prevalence ratio, point estimate (95% CI) | P-value |
| --- | --- | --- |
| **<u>Patient characteristics</u>** |  |  |
| Age, per 1-year increase | 0.995 (0.986 to 1.004) | 0.266 |
| Men vs. Women | 1.08 (0.89 to 1.31) | 0.445 |
| Education level |  |  |
| Junior high school | Reference |  |
| High school | **0.61 (0.44 to 0.83)** | **0.002** |
| University/Graduate school | **0.64 (0.42 to 0.95)** | **0.028** |
| Others | **0.75 (0.57 to 0.996)** | **0.046** |
| Household income |  |  |
| <1,000,000 yen | **0.53 (0.32 to 0.86)** | **0.011** |
| 1,000,000–<5,000,000 yen | 0.74 (0.51 to 1.07) | 0.112 |
| 5,000,000–<10,000,000 yen | 0.85 (0.55 to 1.31) | 0.462 |
| >10,000,000 yen | Reference |  |
| Working |  |  |
| Yes | Reference |  |
| On leave | 0.82 (0.37 to 1.81) | 0.620 |
| No | 1.00 (0.76 to 1.31) | 0.987 |
| Frailty |  |  |
| Non-frailty | Reference |  |
| Very mild to mild frailty | 1.10 (0.81 to 1.49) | 0.559 |
| Moderate to severe frailty | 0.75 (0.37 to 1.54) | 0.436 |
| No. of outpatient visit for RRT selection |  |  |
| None | Reference |  |
| Once | 1.39 (0.96 to 2.04) | 0.085 |
| Twice ore more | **1.59 (1.05 to 2.42)** | **0.029** |
| **<u>Facility characteristics</u>** |  |  |
| Facility type by bed capacity |  |  |
| Clinic (No. of beds <20) | Reference |  |
| Low-capacity hospital (No. of beds 20–<200) | 1.79 (0.85 to 3.76) | 0.125 |
| Low-moderate capacity hospital (No. of beds 200–<400) | 1.51 (0.83 to 2.77) | 0.180 |
| Moderate-to-high capacity hospital (No. of beds ≥400) | **1.99 (1.04 to 3.78)** | **0.037** |
| No. of RRT cases initiated over past one year |  |  |
| <30 cases | Reference |  |
| 30–100 cases | 0.65 (0.42 to 1.01) | 0.054 |
| >100 cases | **0.50 (0.30 to 0.85)** | **0.011** |
| Availability of specialized outpatient visit for RRT selection | 1.17 (0.76 to 1.82) | 0.470 |
| Physicians explaining choice of RRT |  |  |
| Most of the time (70–80% of the time), the physician in charge of specialized outpatient visits for RRT selection / Vary | Reference |  |
| Most of the time (70–80% of the time), the patient's usual nephrologist | 0.90 (0.61 to 1.34) | 0.606 |

<sup>†</sup>Poisson regression with cluster-robust variance using the facilities as cluster units was fitted, including all variables listed above.
RRT, renal replacement therapy; SDM, shared decision-making

Facility-level characteristics also impacted perceived SDM. Patients at moderate-to-high capacity hospitals (≥400 beds) were more likely to perceive RRT selection as involving SDM than those at clinics with fewer than 20 beds (PR: 1.99, 95% CI: 1.04 to 3.78). However, patients at facilities with more than 100 RRT initiations per year were less likely to perceive RRT selection as involving SDM than those at facilities with fewer than 30 initiations (PR: 0.50, 95% CI: 0.30–0.85). The availability of specialized outpatient visits for RRT selection and the type of physician discussing RRT options with patients were not associated with perceptions of SDM.

## Discussion

To the best of our knowledge, this is the first nationwide, multicenter survey in Japan to explore SDM perceptions and preferences for RRT selection among patients with CKD. The survey demonstrated a high percentage of patients perceiving their RRT selection process as involving SDM. It also highlighted key characteristics of the SDM process, including preferred dialogue content, timing and frequency of discussions, desired involvement of specific healthcare professionals, and system-level factors associated with SDM perceptions.

The study revealed that while over 80% of patients perceived their RRT selection process as involving SDM, approximately 80% did not have a clear understanding of SDM prior to making their RRT choice. This significant gap underscores a critical issue and emphasizes the need for enhanced communication and education during routine CKD visits.

The fact that most patients prioritize information about how RRT will affect their daily lives—such as limitations on daily activities and the financial burdens associated with treatment—highlights a gap between what patients seek and what healthcare providers, particularly nephrologists, typically offer.^2,15^ Although discussing the medical aspects of different RRT modalities and prognoses is undoubtedly important, it represents only one of many concerns for patients in their daily lives.^2^ In fact, for older adults with advanced CKD, maintaining independence often takes precedence over survival.^15^ Therefore, the International Society for Peritoneal Dialysis’s recommendation to plan treatment in alignment with the patient’s life goals—while considering the lifestyle of the patient and family—could be applied earlier in the RRT decision-making process. ^16^ Although the JSDT proposals also emphasize the importance of discussing the impact of RRT on daily life as part of appropriate information provision, ^17^ this study suggests that the financial implications should be explicitly included as part of the standard topics for discussion.

Many patients expressed a preference for the timing of SDM to coincide with when RRT became an immediate and tangible concern in their daily lives. Specifically, approximately one-third of patients preferred SDM to occur 6 months to 1 year before RRT initiation, while about 30% preferred it 1 to 3 years prior to RRT initiation. The JSDT recommends providing information about RRT when eGFR falls below 30 mL/min/1.73 m². ^17^ However, a U.S. study estimated the median time to ESKD from an eGFR of 30 mL/min/1.73 m² to be approximately 6 to 10 years.^18^ A U.S. qualitative study highlighted a tendency among patients to focus primarily on living in the “present” rather than planning for the “future.”^2^ Similarly, a U.K. qualitative study suggested that decision-making discussions may feel irrelevant to patients who do not anticipate needing to make decisions for several years.^19^ These findings indicate that the timing of SDM may need to be delayed beyond the point recommended by the JSDT, aligning instead with a timeframe that feels more immediately relevant to patients.

The high priority that patients place on discussing the burden on family and friends may reflect the concept of relational autonomy prevalent in Asia, including Japan.^4^ This emphasis not only highlights a cultural context where family members are often actively involved in medical decision-making, but also underscores the patients’ own prioritization of burden on family. This prioritization stems from deeply rooted values of family obligations, solidarity, and harmony.^20^ However, there are concerns about complex situations where family involvement may inadvertently lead to decisions that overlook the patient’s true wishes owing to implicit pressure.^3,8^ To address this, as outlined in the JSDT proposal for SDM, it is essential to gather sufficient information about the patient’s relationship with their family members.^17^ This ensures a deeper understanding of whether the patient’s desire not to burden their family genuinely reflects their own intentions or arises from external pressures.

Several factors may explain why many patients preferred to receive repeated explanations both before and after RRT initiation. First, decision-making in RRT is inherently a multi-stage process.^19^ For instance, patients must navigate a series of decisions, starting with whether to undergo RRT, followed by selecting a modality (hemodialysis or peritoneal dialysis), and, if hemodialysis is chosen, determining the type of vascular access (arteriovenous fistula, graft, or catheter). Expecting patients to make these complex decisions in a single outpatient visit is unrealistic. Second, patients may find it difficult to engage in meaningful dialogue about RRT options because emotional burdens^4^ and hopelessness when confronting the realities of RRT.^4,21^ The desire for repeated explanations may reflect fluctuating emotions patients experience even after reaching a decision. Third, presenting all the information about RRT in a single, condensed session can overwhelm patients. An Australian qualitative study highlights the importance of iterative discussions, as emphasized by a nephrologist who recognized the need to clarify preferences and confirm decisions over time, while avoiding patient overload. ^3^

Finally, it is worth highlighting the association between multiple outpatient visits for RRT selection—featuring multidisciplinary healthcare provider involvement, including nursing staff, and consultations lasting a minimum of 30 min—and enhanced perceptions of SDM. This finding emphasizes the importance of a system-level approach to SDM, incorporating the contributions of various professionals in addition to physicians, particularly in time-constrained settings.^4,8^ One possible explanation for the observed association between moderate-to-high-capacity hospitals and greater rates of SDM perception may lie in their ability to facilitate more diverse multiprofessional involvement in the RRT decision-making process. For instance, in the multiple outpatient visits for RRT selection, nursing staff can provide repeated explanations of RRT options and engage with patients to explore their preferences, values, and needs.^8^ In the moderate-to-high-capacity hospitals, medical social workers can assist in preparing for the SDM process by offering guidance on financial support and access to social services.^8^ The observation that patients perceive SDM more positively during multiple outpatient visits, as opposed to a single visit, aligns with findings in the general population, where more frequent visits are associated with a higher likelihood of recognizing person-centered interactions that include SDM elements. ^22^ Additionally, given that patients often place greater trust in their usual non-nephrologist physicians with whom they have established longer-term relationships, ^23^ incorporating these professionals’ perspectives into RRT selection may prove valuable.^2^ Care continuity provided by multiprofessional healthcare teams, alongside coordination with patients’ usual non-kidney physicians, should be prioritized to facilitate optimal RRT selection decisions.^8^ Additionally, reimbursement policies should account for these practical and patient-centered care approaches to ensure their sustainability and effectiveness.

The strength of this study lies in its multi-center design, encompassing 49 medical facilities with small to high bed capacities across Japan, thereby ensuring broad generalizability within the Japanese context. This design also enabled the assessment of the influence of facility-level structural variations on SDM perception.

However, some limitations must be acknowledged. First, the participating facilities may have been skewed toward moderate-to high-bed-capacity hospitals, as a relatively large proportion of included facilities were those with well-qualified nephrologists known to the authors. This selection bias may partially explain the high percentage of patients who perceived their RRT selection process as involving SDM. Such facilities often have specialized outpatient services for RRT selection and provide opportunities for multidisciplinary healthcare providers to participate, creating ideal conditions for RRT-related SDM. Second, we did not examine the number of eligible patients with CKD who declined to participate in the study. Based on the number of RRT initiations per quarter at each facility and the duration of the survey, we estimate that approximately 1,850 RRTs were initiated across the participating facilities. Although this total includes patients who opted out or were ineligible owing to emergent RRT initiation or conditions like dementia, it is noteworthy that patients with CKD who participated in the study represented roughly 26% of all RRT initiations. Third, the study excluded conservative kidney management (CKM) from its primary focus. For frail patients or those at high risk of mortality, CKM may provide a better quality of life than RRT,^15,17^ underscoring the need for further research in this area. Fourth, SDM perception was assessed via patient self-report rather than actual outpatient records. Thus, even if healthcare providers presented treatment options, explained their characteristics, and elicited patients’ preferences and values, patients might not recall these interactions accurately. However, as our study targeted patients with CKD who had selected an RRT modality but had not yet commenced treatment, we anticipate that most patients could recall the selection process to a reasonable extent.

In summary, this nationwide study revealed that most patients with CKD in Japan perceived their RRT selection process as involving SDM. Patients prioritized discussions about the effects of RRT on daily life, financial burdens, and the impact on family members over medical aspects. They also expressed a preference for involving their usual non-nephrology physician and a multidisciplinary healthcare team in the RRT decision-making process. Additionally, patients preferred that SDM take place when RRT initiation became an imminent concern, occurring over multiple sessions rather than a single visit. Notably, patients who attended multiple outpatient visits for RRT selection, with participation from nurses and with sufficient time being allocated for discussion, were more likely to perceive the process as SDM.

## Data Availability

The data underlying this article will be shared on reasonable request to the corresponding author.

## Statement

The author(s) used ChatGPT during the preparation of this manuscript to enhance clarity and grammar. All content remains original. Following the use of this tool, the author(s) thoroughly reviewed and edited the material and assume full responsibility for the final content of the publication.

## Author contributions

Y.S. and N.K. conceived and designed the study. N.K. performed the statistical analyses. N.K. developed the figures and tables. Y.S. and N.K. drafted and revised the paper. All authors except for T. Toida and N.K. acquired data. All authors interpreted the data, provided intellectual content, and approved the final version of the manuscript.

## Data sharing statement

The data underlying this article will be shared on reasonable request to the corresponding author.

## Acknowledgements

This study was supported by JSPS KAKENHI (grant numbers: JP19KT0021, JP21K10314, and JP22K19690). The funder had no role in study design, data collection and analysis, decision to publish, or preparation of the manuscript.

## Supplementary Material

Supplementary Figure S1; Supplementary Items 1–4

## Reference

1. Whitney SN, McGuire AL, McCullough LB. A typology of shared decision making, informed consent, and simple consent. Ann Intern Med. 2003;140(1):54–59. 10.7326/0003-4819-140-1-200401060-00012

2. House TR, Wightman A, Rosenberg AR, et al. Challenges to Shared Decision Making About Treatment of Advanced CKD: A Qualitative Study of Patients and Clinicians. Am J Kidney Dis. 2022;79(5):657–666.e1. doi:10.1053/j.ajkd.2021.08.021

3. Dahm MR, Raine SE, Slade D, et al. Older patients and dialysis shared decision-making. Insights from an ethnographic discourse analysis of interviews and clinical interactions. Patient Educ Couns. 2024;122(108124):108124. doi:10.1016/j.pec.2023.108124

4. Yu X, Nakayama M, Wu MS, et al. Shared Decision-Making for a Dialysis Modality. Kidney Int Rep. 2022;7(1):15–27. doi:10.1016/j.ekir.2021.10.019

5. Sakurada T, Koitabashi K, Murasawa M, et al. Effects of one-hour discussion on the choice of dialysis modality at the outpatient clinic: A retrospective cohort study using propensity score matching. Ther Apher Dial. 2023;27(3):442–451. doi:10.1111/1744-9987.13941

6. Kohatsu K, Kojima S, Shibagaki Y, et al. Shared decision-making in selecting modality of renal replacement therapy confers better patient prognosis after the initiation of dialysis. Ther Apher Dial. 2025;29(1):34–41 doi:10.1111/1744-9987.14192

7. Ladin K, Lin N, Hahn E, et al. Engagement in decision-making and patient satisfaction: a qualitative study of older patients’ perceptions of dialysis initiation and modality decisions. Nephrol Dial Transplant. 2017;32(8):1394–1401. doi:10.1093/ndt/gfw307

8. Zagt AC, Bos N, Bakker M, et al. A scoping review into the explanations for differences in the degrees of shared decision making experienced by patients. Patient Educ Couns. 2024;118(108030):108030. doi:10.1016/j.pec.2023.108030

9. Sekimoto M, Asai A, Ohnishi M, et al. Patients’ preferences for involvement in treatment decision making in Japan. BMC Fam Pract. 2004;5(1):1. doi:10.1186/1471-2296-5-1

10. Rockwood K, Theou O. Using the Clinical Frailty Scale in Allocating Scarce Health Care Resources. Can Geriatr J. 2020;23(3):210–215. doi:10.5770/cgj.23.463

11. The Japan Geriatrics Society. Clinical Frailty Scale - Japanese. November 2, 2024. Accessed August 23, 2024. https://www.jpn-geriat-soc.or.jp/tool/pdf/tool_14.pdf

12. Lamer A, Laurent G, Pelayo S, et al. Exploring patient path through Sankey diagram: A proof of concept. Stud Health Technol Inform. 2020;270:218–222. doi:10.3233/SHTI200154

13. Zou G. A modified poisson regression approach to prospective studies with binary data. Am J Epidemiol. 2004;159(7):702–706. doi:10.1093/aje/kwh090

14. Hanafusa N, Abe M, Joki N, et al. 2022 Annual Dialysis Data Report, JSDT Renal Data Registry. J Jpn Soc Dial Ther. 2023;56(12):473–536 (In Japanese except for abstract). doi:10.4009/jsdt.56.473

15. Ramer SJ, McCall NN, Robinson-Cohen C, et al. Health outcome priorities of older adults with advanced CKD and concordance with their nephrology providers’ perceptions. J Am Soc Nephrol. 2018;29(12):2870–2878. doi:10.1681/ASN.2018060657

16. Brown EA, Blake PG, Boudville N, et al. International Society for Peritoneal Dialysis practice recommendations: Prescribing high-quality goal-directed peritoneal dialysis. Perit Dial Int. 2020;40(3):244–253. doi:10.1177/0896860819895364

17. Okada K, Tsuchiya K, Sakai K, et al. Shared decision making for the initiation and continuation of dialysis: a proposal from the Japanese Society for Dialysis Therapy. Ren Replace Ther. 2021;7(1). doi:10.1186/s41100-021-00365-5

18. Grams ME, Li L, Greene TH, et al. Estimating time to ESRD using kidney failure risk equations: results from the African American Study of Kidney Disease and Hypertension (AASK). Am J Kidney Dis. 2015;65(3):394–402. doi:10.1053/j.ajkd.2014.07.026

19. Joseph-Williams N, Williams D, Wood F, et al. A descriptive model of shared decision making derived from routine implementation in clinical practice (’Implement-SDM’): Qualitative study. Patient Educ Couns. 2019;102(10):1774–1785. doi:10.1016/j.pec.2019.07.016

20. Alden DL, Friend J, Lee PY, et al. Who Decides: Me or We? Family Involvement in Medical Decision Making in Eastern and Western Countries. Med Decis Making. 2018;38(1):14–25. doi:10.1177/0272989X17715628

21. Kurita N, Wakita T, Ishibashi Y, et al. Association between health-related hope and adherence to prescribed treatment in CKD patients: multicenter cross-sectional study. BMC Nephrol. 2020;21(1):453. doi:10.1186/s12882-020-02120-0

22. Okamura M, Fujimori M, Otsuki A, et al. Patients’ perceptions of patient-centered communication with healthcare providers and associated factors in Japan - The INFORM Study 2020. Patient Educ Couns. 2024;122(108170):108170. doi:10.1016/j.pec.2024.108170

23. Barrett TM, Green JA, Greer RC, et al. Advanced CKD Care and Decision Making: Which Health Care Professionals Do Patients Rely on for CKD Treatment and Advice? Kidney Medicine. 2020;2(5):532–542.e1. doi:10.1016/j.xkme.2020.05.008

