## Supplementary Files for "Shared Decision-Making in Renal Replacement Therapy Selection: Patient Perceptions, Preferences, and Influencing Factors in a Nationwide Cross-Sectional Study in Japan"

**Supplementary Material**

**Supplementary Figure S1. National Map of Participating Facilities**

The zip codes of participating facilities have been mapped using Google My Maps. To view the distribution on Google My Maps, please click the following link: https://tinyurl.com/mzhhx5vu.

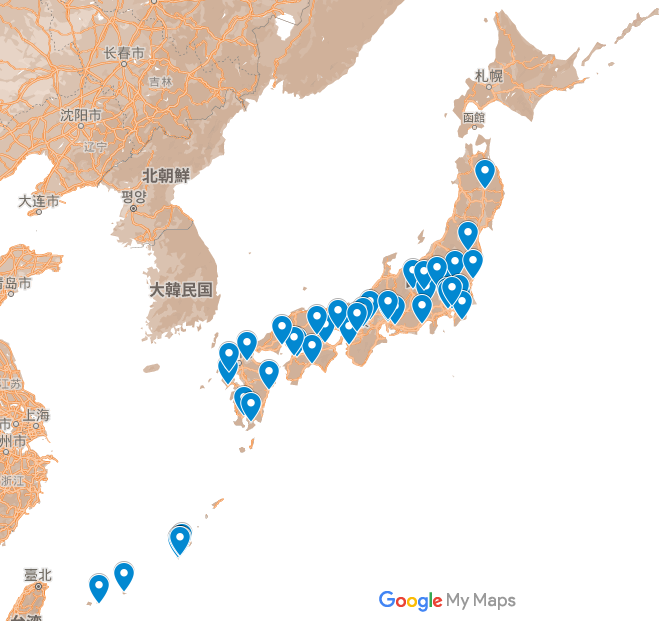

**Supplementary Item S1. Full details of the other members of the PREPARES Study Group^☨^**

^☨^The PREPARES Study stands for **PREference for PAtient REnal replacement therapy and Sharing Study**.

Shigeyuki Arai, MD, PhD^1^; Tsuyoshi Watanabe, MD, PhD^2^; Keita Iwasaki, MD^2^; Yuuki Itou, MD^2^; Fumika Nagase, MD^2^; Kenta Torigoe, MD, PhD^3^; Shinichi Abe, MD, PhD^3^; Kumiko Muta, MD, PhD^3^; Tomomi Endo, MD, PhD^4^; Keita Mori, MD, PhD^4^; Michiya Shinozaki, MD, PhD^5^; Megumi Oikawa, MD, PhD^6^; Tsuyoshi Ohshiro, MD^6^; Yoshitaka Ishibashi, MD, PhD^7^; Ryo Sugiyama, MD^8^;

^1^Division of Nephrology, Department of Internal Medicine, Teikyo University School of Medicine, Itabashi-Ku, Tokyo, Japan

^2^Department of Rheumatology and Nephrology, Chubu Rosai Hospital, Nagoya-City, Aichi, Japan

^3^Department of Nephrology, Nagasaki University Hospital, Nagasaki-City, Nagasaki, Japan

^4^Medical Research Institute Kitano Hospital, PIIF Tazuke-Kofukai, Osaka-city, Osaka, Japan

^5^Department of Nephrology, Shin-Yurigaoka General Hospital, Kawasaki-City, Kanagawa, Japan

^6^Division of Nephrology, Department of Internal Medicine, Showa University Fujigaoka Hospital, Yokohama-City, Kanagawa, Japan

^7^Department of Nephrology, Japanese Red Cross Medical Center, Shibuya-ku, Tokyo, Japan

^8^Department of internal medicine, Okinawa prefectural Yaeyama Hospital, Ishigaki-city, Okinawa, Japan

**Supplementary Item S2. Questionnaire Items Regarding the Choice of Renal Replacement Therapy**

1. Was your current primary kidney doctor involved in your choice of renal replacement therapy?

Yes

No

2. Have you and your kidney doctor ever discussed the following? (Please circle all that apply.)

1. Whether or not to pursue renal replacement therapy (dialysis or kidney transplant).

2. Which type of renal replacement therapy to choose (hemodialysis, peritoneal dialysis, or kidney transplant).

3. Whether to proceed with or not yet proceed with medical preparation for the selected renal replacement therapy (such as vascular access surgery for hemodialysis or catheter placement for peritoneal dialysis).

4. Whether to start or not yet start the selected renal replacement therapy in the near future.

5. Other (please specify): _______________

**Supplementary Item S3. Introductory Statements for the Questionnaire on Shared Decision-Making**

*Please read this page carefully before proceeding to the questions on the next page.*

Renal replacement therapy options include hemodialysis, peritoneal dialysis, and kidney transplantation, and you will need to choose the one that best suits you.

Choosing the right treatment depends not only on your medical condition but also on your personal values, outlook on life, and lifestyle, including your daily schedule and activities. Therefore, it is important to consider not only the medical suitability of each option but also what matters most to you personally, such as your values, hobbies, preferences, and lifestyle.

We recommend discussing these options with your healthcare providers (e.g., doctors, nurses) and trusted individuals (e.g., family members, close friends) to ensure a comprehensive understanding and to choose the option that feels right for you. This process of collaborative discussion is called Shared Decision-Making (SDM).

SDM differs from a process in which treatment options are chosen based solely on the opinion of healthcare providers or only on the preferences of the patient or family members.

In SDM, everyone shares insights on your living environment, lifestyle, hobbies, preferences, and values, along with considering medical factors, to identify which treatment option is most suitable to you. Through these discussions, healthcare providers and patients work together to reach a consensus on the best treatment plan.

Since deciding on a treatment option can have a significant impact on your life, SDM discussions may take more than one session. Sometimes, it may take several sessions to reach a consensus, or, occasionally, a decision may be postponed if consensus is not reached. Preparing for renal replacement therapy typically requires several months, and often up to six months. Additionally, even if you reach an initial agreement on a treatment, you can always revisit and change your decision later if you change your mind during the course of treatment.

In SDM discussions, the healthcare team may include professionals beyond just your doctor, such as nurses and other specialists. They can often share unique insights regarding the treatment's impact on your life. On your side, you may invite a key person, such as a family member or close friend who understands your life, personality, and values, to help you think through the decision. This can be a great support in considering which treatment option best aligns with your values, lifestyle, and family situation. Ultimately, however, the final decision is yours.

**Supplementary Item S4. Questionnaire Items Regarding Shared Decision-Making**

1. Were you aware of shared decision-making (SDM) as a way to choose your renal replacement therapy (e.g., dialysis or transplant)? (Please select one.)

1. I am well aware of it.

2. I am somewhat aware of it.

3. I have heard of it, but I am not very familiar with it.

4. I am not at all aware of it.

2. Do you feel that your renal replacement therapy was chosen through the SDM approach? (Please select one.)

1. Strongly agree

2. Somewhat agree

3. Somewhat disagree

4. Strongly disagree

Please answer the following questions as if you were choosing your renal replacement therapy (dialysis or transplantation) through shared decision-making (SDM).

(1) When do you think SDM should begin? (Please select one.)

*... before starting renal replacement therapy:*

a. More than 5 years before

b. 3 to 5 years before

c. 1 to 3 years before

d. 6 months to 1 year before

e. Less than 6 months before

(2) Would you like your usual kidney doctor to participate in SDM for choosing your renal replacement therapy? (Please select one.)

1. Agree

2. Neither agree nor disagree

3. Disagree

(3) Please select other healthcare providers you would like to participate in SDM. (Please select all that apply.)

1. Healthcare providers with expertise in renal replacement therapy (e.g., nurses, clinical engineers, transplant coordinators)

2. Social workers (medical social workers)

3. Usual doctor (not specialized in kidney care)

4. Primary home care nurse

5. Care manager

6. Others (please specify): ______

(4) During SDM, what additional information would you like to ask your healthcare provider about, beyond the specific treatment methods? (Please select all that apply.)

1. Impact of treatment on family and friends

2. Impact of treatment on social activities (e.g., socializing, clubs)

3. Life expectancy without dialysis or transplantation

4. Ability to continue fulfilling household roles with each treatment

5. Impact of treatment on exercise, hobbies, and travel

6. Ability to continue working after treatment

7. Average life expectancy after starting each treatment

8. Daily life limitations with each treatment

9. Economic burden (financial impact) of each treatment

(5) The choice of renal replacement therapy can significantly impact your life, so we will dedicate time during the shared decision-making (SDM) process to discuss it. This discussion can happen more than once. How often should SDM discussions occur? (Please select one.)

a. About once a year

b. About once every six months

c. Once every few months

d. Every visit

e. As needed

f. Just once

g. Other (please specify): ______

(6) Even after you have chosen renal replacement therapy, you may want to change your treatment due to changes in your physical condition or feelings. In that case, you can still modify your treatment. After you and your healthcare provider have selected your renal replacement therapy, how often should we check in to see if you would like to change your treatment? (Please select one.)

a. About once a year

b. About once every six months

c. Once every few months

d. Every visit

e. As needed

f. Not necessary

g. Other (please specify): ______

(7) Even after starting renal replacement therapy, it may be possible to change to other treatment methods. What are your thoughts on performing shared decision-making (SDM) for this purpose? (Please select one)

1. I would like to have SDM after starting renal replacement therapy.

2. I do not need SDM after starting renal replacement therapy.

3. I am unsure.
